# Angiotensin II Regulates Anxiety and Social-Affective Top-Down and Bottom-Up Attention Control in a Sex-dependent Manner

**DOI:** 10.1101/2025.10.26.25338807

**Authors:** Mengfan Han, Kun Fu, Wenyi Dong, Junjie Wang, Dan Liu, Qian Zhuang, Xiaolei Xu, Ferraro Stefania, Ting Xu, Keith M Kendrick, Dezhong Yao, Wing Chung Chang, Benjamin Becker

**Affiliations:** The Center of Psychosomatic Medicine, Sichuan Provincial Center for Mental Health, Sichuan Provincial People’s Hospital, University of Electronic Science and Technology of China; MOE Key Laboratory for Neuroinformation, School of Life Science and Technology, University of Electronic Science and Technology of China, Chengdu, China; Center for Cognition and Brain Disorders, The Affiliated Hospital of Hangzhou Normal University, Hangzhou, Zhejiang, China; Faculty of Psychology, Shandong Normal University, Jinan, China; Faculty of Psychology, Southwest University, Chongqing, China; Department of Psychiatry, School of Clinical Medicine, University of Hong Kong, Hong Kong SAR, China; Department of Psychology, The University of Hong Kong, Hong Kong SAR, China

## Abstract

The renin-angiotensin system (RAS), traditionally known for cardiovascular regulation, has increasingly been recognized as a modulator of cognitive and affective functions. However, whether the RAS regulates attentional control and whether such effects are sex-dependent remain unexplored.

The present preregistered, randomized, double-blind, placebo-controlled pharmacological eye-tracking study (N = 79) examined the effects of transient angiotensin II type 1 receptor (AT1R) blockade via losartan (50 mg) on emotional attention control using a validated anti-saccade task with social (emotional faces) and non-social stimuli. Treatment effects on state anxiety and oculomotor responses were characterized using traditional performance metrics and a novel trial-history informed dynamic control (TIDC) framework for adaptive control.

Losartan reduced state anxiety irrespective of sex and induced sexually dimorphic reconfiguration of attentional processing. In females, AT1R blockade enhanced performance by reducing endpoint error without altering latency. Conversely, in males, losartan increased endpoint error and prolonged latency of the first correct saccade. Trial-history analyses further revealed that losartan reduced error probabilities following error and repeat trials in both sexes. Yet, following correct trials, females receiving losartan maintained lower error probabilities, while males receiving losartan exhibited higher errors, potentially reflecting a failure to flexibly disengage from the effortful controlled mode.

Findings indicate that the RAS modulates anxiety and attentional control, the latter in a sex- dependent manner. AT1R blockade can reconfigure attentional processing and adaptive control, suggesting sex-specific therapeutic potential in disorders characterized by excessive anxiety and attentional dysregulation.

## 1 Introduction

The flexible regulation of attention is fundamental to adaptive behavior, requiring the suppression of salient yet irrelevant stimuli while focusing on task-relevant information (Anderson, 2021; Oberauer, 2024). This capacity, referred to as attentional control, is a multidimensional construct that encompasses both top-down, goal-directed regulation of attention and bottom-up, stimulus-driven attentional capture (Pinto et al., 2013). A core subcomponent of this system is inhibitory control, which specifically refers to the active suppression of prepotent but inappropriate responses or mental representations (Munoz & Everling, 2004). In anxiety and trauma-related disorders, deficits in these control processes are considered a central pathological cognitive mechanism, characterized by an imbalance where bottom-up salience processing overrides topdown goal-directed processing. This imbalance manifests as excessive attention to threat-related information (Boal et al., 2018; Sussman et al., 2016). Consequently, interventions aimed at improving attentional and inhibitory control represent a crucial therapeutic candidate therapy for these conditions (e.g. Li et al., 2020).

Current first-line treatments for anxiety, such as SSRIs and cognitive-behavioral therapy, have limited efficacy or delayed onset in a subset of patients (Bandelow et al., 2017; Capron et al., 2017), underscoring the need for novel interventions. Beyond traditional monoaminergic systems, the renin-angiotensin system (RAS) has recently attracted attention for its role in emotion and cognitive regulation (Swiercz et al., 2020; Xu et al., 2024, 2025). The central RAS primarily acts through the angiotensin II type 1 receptor (AT1R), and pharmacological blockade of this receptor (e.g., via losartan) has been shown to enhance fear extinction and reduce anxiety-like behaviors in preclinical models (Chrissobolis et al., 2020; Gao et al., 2021; Swiercz et al., 2020; Yang et al., 2023). Recent translational studies extended these findings to humans and indicate that a single dose of an AT1R antagonist can reduce amygdala reactivity in high-trait anxiety individuals and strengthen prefrontal control over the amygdala, thereby improving fear regulation (Prasad et al., 2025; Xu et al., 2022; Zhou et al., 2019).

Mechanistically, this improvement in regulation is thought to rely on AT1R-mediated modulation of prefrontal cortex (PFC) function. Preclinical evidence suggests that angiotensin II may exert an inhibitory influence via prefrontal AT1R and (partly) via dopaminergic modulation (Kobiec et al., 2021; Ott & Nieder, 2019). This mechanism is underscored by human neuroimaging evidence showing that AT1R blockade enhances activity and connectivity within frontocingulate control circuits— including the dorsolateral PFC and anterior cingulate cortex— during tasks of cognitive and emotional regulation (e.g., T. Xu et al., 2023, 2024; F. Zhou et al., 2019). Based on this, we hypothesize that AT1R blockade will enhance top-down cognitive control, encompassing both inhibitory control and goal-directed execution.

To assess these distinct components we combined the transient pharmacological blockade of the AT1R with an emotional pro- and anti-saccade task (Xu et al., 2019; Zhuang et al., 2021). This paradigm effectively dissociates the sub-processes of control: the pro-saccade task measures bottom-up, reflexive attentional orienting, while the anti-saccade task requires the suppression of this reflex alongside the execution of a volitional saccade (top-down control), processes reliant on the dorsolateral prefrontal cortex (DLPFC) and anterior cingulate cortex (ACC) (Cieslik et al., 2016; Munoz & Everling, 2004; Si et al., 2022). Accordingly, we used directional error rate to index inhibition failure, while metrics from correct trials characterized the quality of volitional execution. Specifically, saccadic latency quantifies the speed of sensorimotor processing and motor planning, and endpoint error assesses the spatial accuracy of the saccade (Kanaan & Moacdieh, 2022; Noorani & Carpenter, 2016).

Beyond these conventional measures, emerging evidence indicates that AT1R may additionally modulate overarching behavioral adaptations, particularly in response to threat (Zika et al., 2024) and negative feedback (Xu et al., 2023). Classical analyses of the anti-saccade task focus on condition-level performance (e.g., average accuracy); however, sequential adaptations — such as post-error slowing (PES), post-error improvement in accuracy (PIA), and stimulus switch effects— provide critical insights into the trial-by-trial dynamics of error monitoring and cognitive flexibility (Monsell, 2003; Notebaert et al., 2009). Surprisingly, despite these effects being extensively studied in cognitive control, no study has systematically examined them within an emotional anti-saccade context, nor explored how pharmacological interventions impact these dynamic processes. To address this, we developed a trial-history informed dynamic control framework (TIDC) to capture these adjustments and examine whether AT1R modulates the corresponding adaptation.

Finally, despite long-standing evidence that AT1R expression and signaling differ between sexes (Bold & L, 1999; Chen et al., 1992), previous studies have often overlooked or failed to find sex differences in the effects of AT1R blockade on social-cognitive functions (Prasad et al., 2025; Zhou et al., 2023). Given that males typically exhibit higher AT1R density (Bachmann et al., 1991) and dopaminergic regulation shows sexual dimorphism (Sacher et al., 2013), it is biologically plausible that AT1R blockade exerts sex-specific effects on attentional control. Exploring this is crucial for understanding individualized treatment responses.

Against this background, the present pre-registered, randomized, placebo-controlled eyetracking study aimed to explore the effects of a single dose of a selective AT1R antagonist (50mg Losartan) on bottom-up and top-down attentional control. We placed particular emphasis on social- emotional processing, trial-by-trial behavioral adaptation, and potential sex differences. Employing a validated emotional anti-saccade task with our innovative TIDC analysis, we hypothesized that transient AT1R blockade would: (1) improve anti-saccade performance, reflecting enhanced topdown attentional control; (2) facilitate both post-error adjustment and stimulus-switch processing, reflecting improved behavioral adaptation; and (3) exert different effects on attentional control and behavioral adaptation between males and females.

## 2 Method

### 2.1 Participants

A total of 79 healthy participants were recruited for the preregistered pharmacological eyetracking experiment (losartan [LT], n=39; placebo [PLC], n=40; see CONSORT flowchart for details in **Fig. 1**). This sample size exceeded the requirement of 52 participants determined by an a priori G+Power analysis (effect size *f* = 0.20, power = 80%, α = 0.05) for a 2 (Treatment) 2 (Treatment) × 2 (Sex) × 2 (Task) × 2 (Condition) mixed-design ANOVA, thus ensuring adequate statistical power.

The study was approved by the local ethics committee of University of Electronic Science and Technology of China (UESTC), conducted according to the Declaration of Helsinki, and preregistered (ClinicalTrials.gov ID: NCT06329050). All participants provided written informed consent and received monetary compensation (140 RMB).

**Fig 1.**
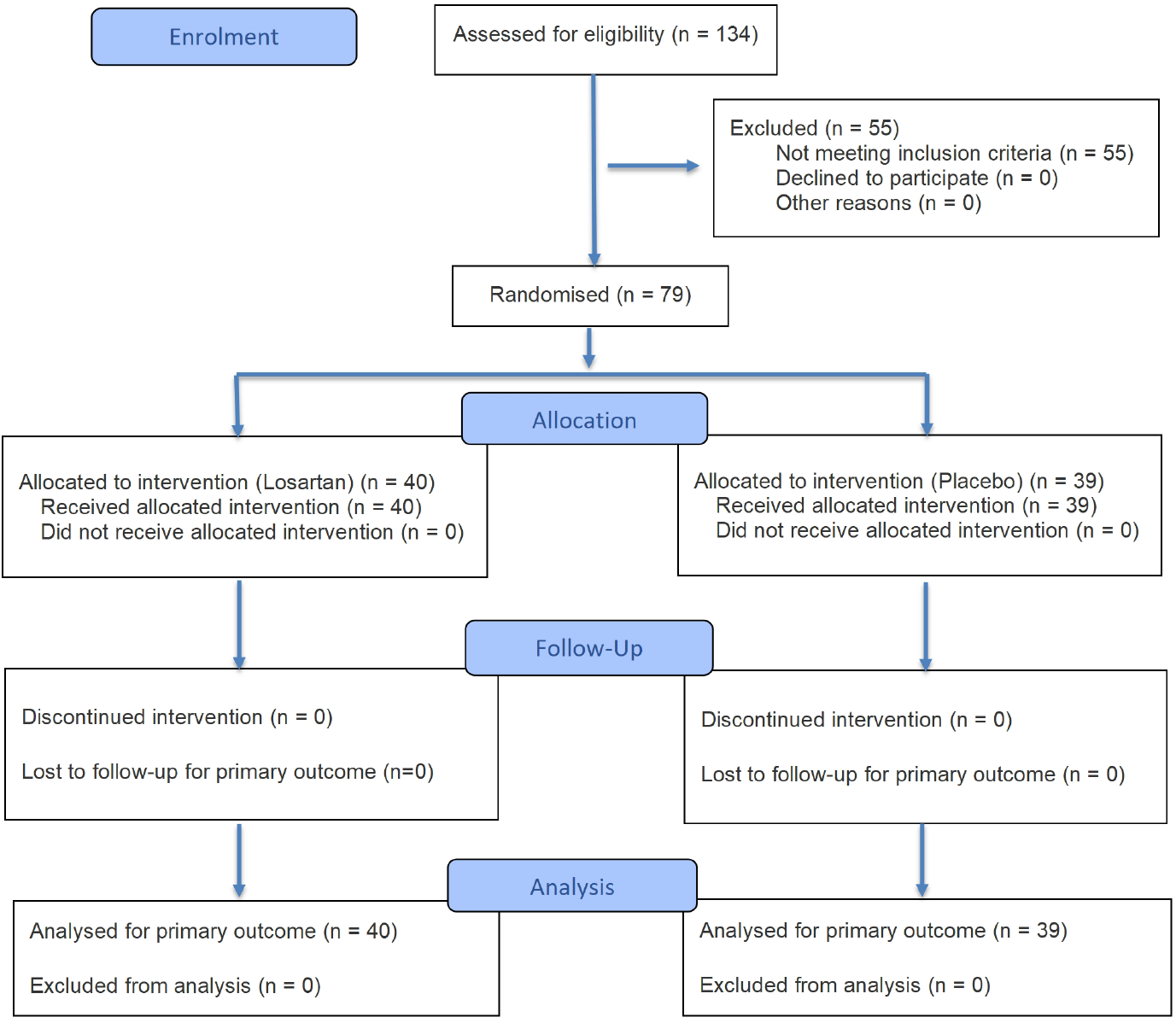
CONSORT flowchart. Detailed exclusion criteria are provided in the supplementary materials.

### 2.2 Study Protocols

We employed a randomized, double-blind, placebo-controlled, between-subject design to examine neurocognitive effects of transient AT1R blockade using a pro- and anti-saccade tasks (**Fig. 2A**). Participants were randomly assigned to receive either a single 50-mg dose of LT or PLC, administered orally in identical capsules. Randomization was computer-generated and implemented by an independent researcher who had no direct contact with participants or experimenters.

Mood and anxiety assessments were conducted using validated Chinese versions of standardized questionnaires. Prior to drug administration, participants completed the Positive and Negative Affect Schedule (PANAS) (Watson et al., 1988), the State-Trait Anxiety Inventory (STAI) (Spielberger, 2012), and the Liebowitz Social Anxiety Scale (LSAS) (Heimberg et al., 1999) to assess baseline affective states and fear-related traits. State anxiety (STAI state subscale) was reassessed immediately after the experiment. These questionnaires were administered to verify baseline comparability between groups.

Cardiovascular measurements (blood pressure and heart rate) were taken at baseline, peak drug effect (90 minutes post-administration), and post-experiment. In line with the pharmacokinetics of LT - peak plasma levels occurring at approximately 90 minutes and a terminal elimination half-life of 1.5-2.5 hours (Collister et al., 2002; Sica et al., 2005; Xu et al., 2022) - all experimental procedures were timed to commence during peak plasma levels.

### 2.3 Paradigm

The present study employed a validated anti-saccade paradigm (Chen et al., 2014; Zhuang et al., 2022), featuring five social-emotional facial expressions (angry, sad, happy, fearful, and neutral) from eight actors (4 male, 4 female) and eight non-social oval shapes with slight variations. To prevent affective carry-over effects, the task began with two non-social blocks (one anti-saccade and one pro-saccade block), each containing 48 trials. These were followed by 12 emotional blocks (6 anti-saccade and 6 pro-saccade blocks), with 40 trials per block (8 trials per emotion). Block order (anti- vs. pro-saccade) and trial sequence within blocks were randomized.

Each block started with a 2000ms instructional cue (“Towards” or “Away”; **Fig. 2B**), followed by a jittered fixation period (mean duration: 1500ms; range: 1000–3500ms), during which participants were instructed to maintain fixation. Subsequently, a stimulus appeared on either the left or right side of the screen for 1000ms. In “Away” blocks (anti-saccade task), participants were required to look in the direction opposite to the stimulus, while in “Towards” blocks (pro-saccade task), they were instructed to look toward the stimulus as quickly and accurately as possible. The task lasted approximately 40 minutes, with brief rest periods provided between blocks.

**Fig 2.**
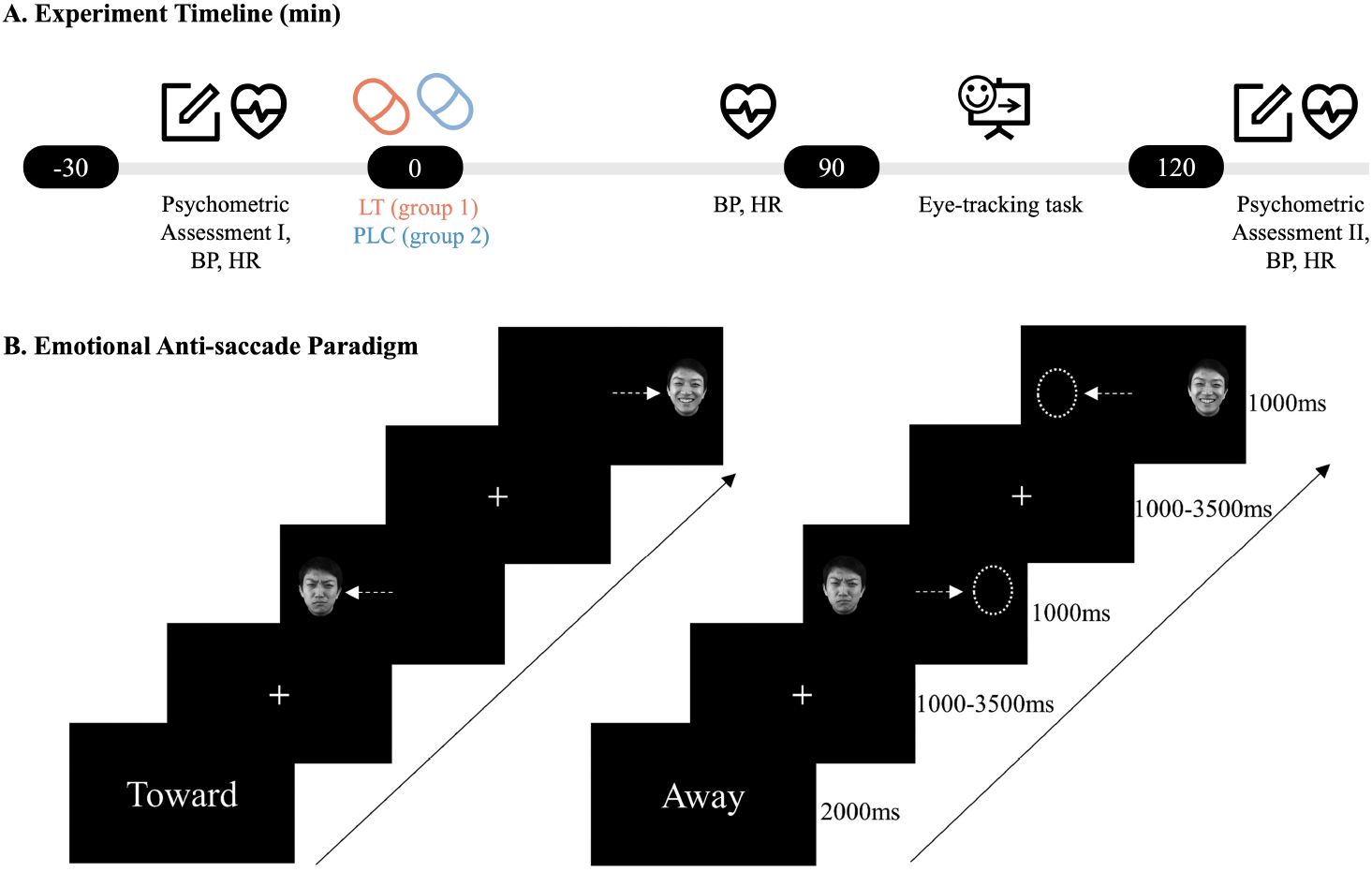
**(A)** Experimental timeline showing the measurement points and drug administration schedule. **(B)** The emotional anti-saccade paradigm and timing set including the pro-saccade (left) and anti-saccade task (right).

### 2.4 Data recording and processing

Eye movement data were recorded using an EyeLink 1000 Plus system (SR Research, Ottawa, Canada) sampling at 2000 Hz with a screen resolution of 1024 × 768. A chin rest positioned 57 cm from the monitor ensured standardized viewing distance and position. A 9-point calibration was performed before each block to ensure eye-tracking data quality.

Raw eye-tracking data were preprocessed and exported using EyeLink DataViewer 3.1 (SR Research, Mississauga, Ontario, Canada). Following established procedures in saccade paradigms (Antoniades et al., 2013; Hutton & Ettinger, 2006; Xu et al., 2019; Zhuang et al., 2021), trials were excluded based on the following criteria: latencies < 70 ms or > 700 ms, saccade velocities below 30°/sec, or initial fixation deviations exceeding 2°.

### 2.5 Statistical Analyses

#### 2.5.1 Initial Saccade Analyses

We analyzed initial saccades using a hierarchical approach based on established oculomotor control paradigms (Hutton & Ettinger, 2006; Luna et al., 2008). First, we examined inhibitory control efficiency through directional error rates (proportion of initial saccades opposite to the target direction) using a 2 (Treatment) × 2 (Sex) × 2 (Task) × 2 (Condition) repeated measures ANOVA. Next, to assess the quality of volitional motor planning, we analyzed both latency (time from stimulus onset to saccade initiation) and endpoint precision (Euclidean distance from landing position to target center) of the first correctly executed saccade using the same factorial design. Finally, to probe emotion-specific modulation of these control processes, we conducted a follow-up ANOVA replacing the binary Condition with six Stimulus types (angry, fear, sad, happy, neutral, shape) in a 2 (Treatment) × 2 (Sex) × 2 (Task) × 6 (Stimulus) design. This three-step procedure systematically evaluated performance from basic task compliance to motor execution and finally to emotional specificity.

#### 2.5.2 Trial-history informed dynamic control (TIDC) Analyses

We analyzed trial-history effects exclusively within emotional face blocks to ensure that both stimulus switching and post-error adjustments were assessed within the domain of social information processing, thereby capturing control dynamics intrinsic to this context (Adolphs, 2009; Kanwisher et al., 1997). We focused on two key markers that capture the moment-to-moment regulation of the cognitive control system:

1. **Post-error adjustments** (Prev_Error: correct vs. error on trial n-1). This comparison isolates the recruitment of reactive control mechanisms triggered by performance failure (post-error trials) relative to the successful performance (post-correct trials), indexing the system’s capacity for moment-to-moment behavioral regulation (Danielmeier & Ullsperger, 2011).
2. **Emotion-switching costs** (Stimulus_Switch: repetition vs. switch of stimulus type from trial n-1). This contrast isolates the cognitive cost of reconfiguring attentional sets (in switch trials) relative to maintaining the current task set (in repetition trials), thereby serving as a robust index of cognitive flexibility (Monsell, 2003).

Together, these measures operationalize trial-history informed dynamic control by quantifying how prior trial outcomes and stimulus history modulate current behavior.

Generalized linear mixed models (GLMMs for error probability) and linear mixed models (LMMs for latency) are used to examine these effects. The maximal model for each outcome variable included fixed effects for Task, Treatment, Sex, Prev_Error, and Stimulus_Switch, encompassing all possible two-way and three-way interactions between these factors. The model structure was specified as:

Current_Error/Latency ∼ Task × Treatment × Sex × Prev_Error × Stimulus_Switch + (1 + Prev_Error × Stimulus_Switch || Subject)

This specification explicitly excludes higher-order (four-way and five-way) interactions for reasons of parsimony and interpretability (Brauer & Curtin, 2018; Meteyard & Davies, 2020). Random effects included random intercepts and uncorrelated slopes for the sequential effects of Prev_Error, Stimulus_Switch, and their interaction (Barr et al., 2013).

#### 2.5.3 Software and Statistical Reporting

Statistical analyses were performed using SPSS 26 for ANOVAs and Python 3.12.5 (utilizing R’s lme4 package via rpy2 interface) for mixed models (GLMM/LMM). The False Discovery Rate (FDR) procedure (*q* = 0.05) was applied to correct for multiple comparisons. Results are reported as partial eta-squared (*η*^2p^) for ANOVAs, odds ratios (OR) with 95% CIs for GLMMs, and standardized coefficients (*β*) for LMMs.

## 3 Results

### 3.1 Potential Confounders

#### 3.1.1 Demographics, effects on anxiety and cardiovascular parameters

The LT and PLC groups showed no differences in age, mood, or personality traits at baseline (all *p*s > .05; see Table S1 for questionnaire scores). Examining effects of LT on anxiety using a 2 (Treatment: LT vs. PLC) × 2 (Sex: male, female) × 2 (Time: pre vs. post) mixed ANOVA on State Anxiety revealed significant Treatment × Time interaction (*F* = 4.35, *p* = .04, *η*^2p^ = 0.056) (**Fig 3A**). Between-group comparison showed lower post-task anxiety scores in the LT group compared to PLC (*p* = .014, *d* = 0.60). Within-group analysis revealed significant anxiety reduction in the LT group (pre: 37.51 ± 1.10, post: 33.41 ± 1.34, *p* = .004, *d*z = 0.49) but not in the PLC group (pre: 38.17 ± 1.07, post: 38.08 ± 1.30, *p* = .939, *d*z = 0.02), suggesting an anxiolytic effect of LT. No main or interaction effects of sex were observed.

Examining cardiovascular effects using separate 2 (Treatment) × 2 (Sex) × 3 (Time: baseline, drug peak effect [80 minutes post-LT], post-task) mixed ANOVAs were conducted for systolic/diastolic blood pressure [SBP/DBP] and heart rate. Consistent with previous findings (Franklin et al., 1997), males exhibited higher resting blood pressure than females (SBP: 117.68 ± 0.88 vs. 110.57 ± 0.90 mmHg, *p* < .001; DBP: 74.58 ± 1.41 vs. 71.29 ± 1.06 mmHg, *p* = .031) (**Fig 3B**). Critically, a single dose of LT did not produce a significant blood pressure-lowering effect compared to PLC at any time point (*p*s > .10; see Table S2), which aligns with our previous studies and reflects the delayed peak cardiovascular efficacy of losartan in normotensive individuals (Reinecke et al., 2018; Xu et al., 2022, 2023).

### 3.2 Initial Saccade Performance

#### 3.2.1 Direction Error Rate

Analysis revealed the expected main effects of Task (*F* = 132.13, *p* < .001, *η*^2p^ = 0.64) and Condition (*F* = 9.04, *p* = .004, *η*^2p^ = 0.11), with higher error rates in the anti-saccade compared to the pro-saccade task and in social versus nonsocial condition. This pattern replicates established findings (Xu et al., 2019; Zhuang et al., 2022) and validates the effectiveness of our experimental manipulation. However, no significant effects of Treatment or Sex were observed (all *p*s > .05; complete statistics are provided in the Supplementary Materials).

#### 3.2.2 Endpoint Error

A significant four-way interaction between Treatment, Sex, Task, and Condition (*F* = 5.74, *p*=.019, *η*^2p^=0.073) was observed on endpoint error (**Fig 3C**). In the anti-saccade task under nonsocial conditions, LT males demonstrated significantly larger endpoint errors than PLC males (160.07px vs 111.84px, *p* = .004), while females showed a marginal opposite trend (108.27px vs 131.54px, *p* = .056). Subsequent between-sex comparisons revealed that males in the LT group exhibited greater deviation than females (*p* = .004), whereas this pattern was reversed in the PLC group (*p* = .061). No other Task × Condition combinations showed significant treatment or sex effects (all *p*s > .26). Additional follow-up analyses that replaced the binary Condition with the six specific stimulus types did not reveal any further significant effects beyond the primary social/nonsocial distinction.

#### 3.2.3 Latency

Analysis revealed a significant Treatment × Condition interaction (*F* = 7.51, *p* = .008, *η*^2p^ = 0.09), with the LT group showing longer latencies than PLC in non-social conditions (*p*=.013) but not in social conditions (*p* = .59; **Fig. 3D**). Follow up 2 (Treatment) × 2 (Sex) × 2 (Task) × 6 (Stimulus) analysis revealed a significant Treatment × Sex × Stimulus interaction (*F* = 4.32, *p* = .002, *η*^2p^ = 0.23) where LT males exhibited longer latencies specifically for shape stimuli compared to PLC males (*p* = .035), while no treatment differences were observed in females (*p* = .148).

**Fig 3.**
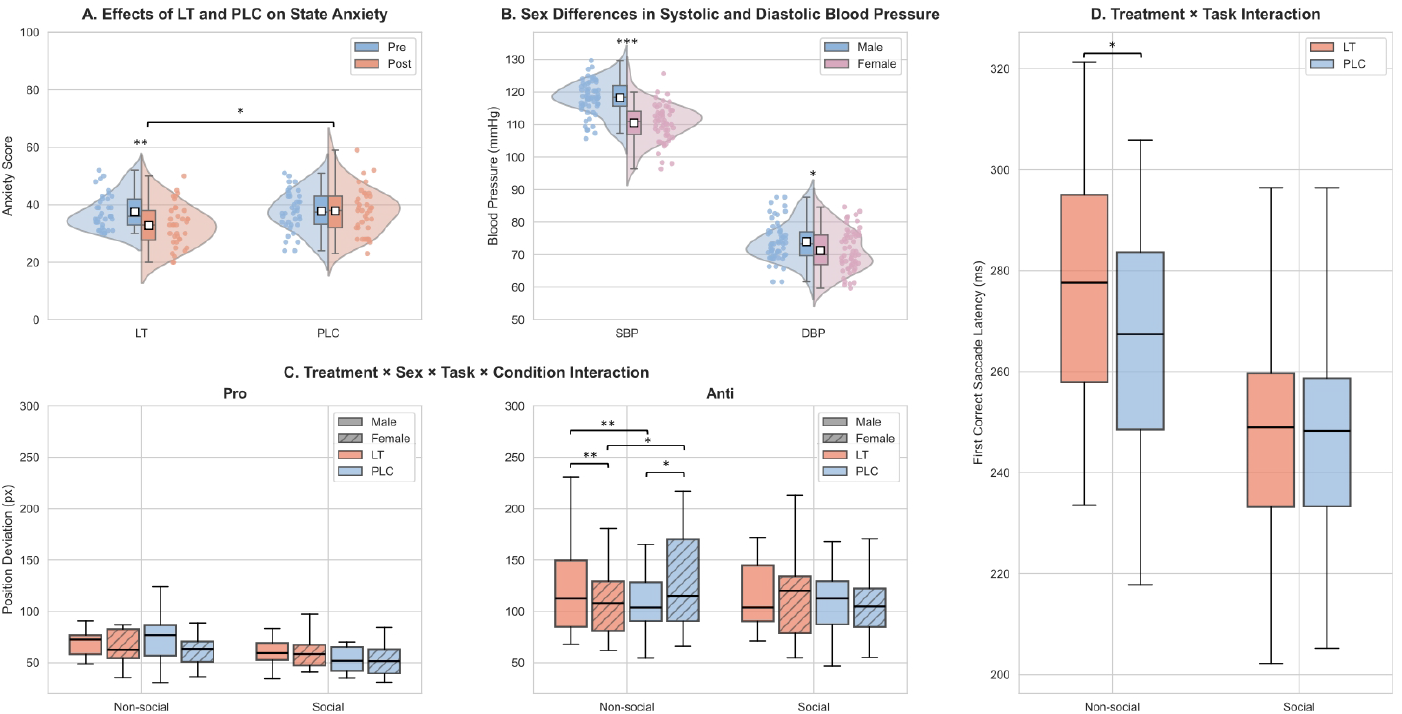
Assessment of Potential Confounders and Initial Saccade Performance. **(A) State anxiety scores** (STAI-S) before and after the experiment in LT and PLC groups; **(B) Systolic and diastolic blood pressure** (mmHg) by sex. Data presented as raincloud plots (individual data points, probability density distributions, and box plots with quartiles and median). **(C) Endpoint precision** (pixels) in pro-saccade (left) and antisaccade tasks (right). Pro-saccade performance showed no significant group differences. In anti-saccade under non-social conditions, males showed larger endpoint error under LT versus PLC, while females displayed the opposite pattern. Sex differences within treatments: higher endpoint error in females versus males under PLC, but lower under LT. **(D) Saccadic latency** (ms) was longer in LT versus PLC specifically under non-social conditions, with no group differences in social condition. Data in C and D shown as box plots (median, IQR, whiskers at 1.5 × IQR). Abbreviations: LT, Losartan; PLC, placebo; STAI-S, State-Trait Anxiety Inventory-State; IQR, interquartile range. ^*^*p* < .05, ^**^*p* < .01, ^***^*p* < .001

### 3.3 Trial-history Dependent Saccade Control

#### 3.3.1 Current-trial Error Probability Modulation

Significant main effects were observed across all factors. Error probability was lower in the LT group compared to PLC (OR = 0.86, 95%CI [0.77, 0.96], *p* = .008), in males compared to females (OR = 0.84, 95%CI [0.75, 0.94], *p* = .003), and in pro-saccade tasks compared to anti-saccade (OR = 2.09, 95%CI [1.86, 2.34], *p* < .001). Trial history also exerted strong effects: error rates were lower on switch trials (vs. repeat; OR = 0.10, 95%CI [0.08, 0.13], *p* < .001) and markedly reduced following errors (vs. prev_correct; OR = 0.24, 95%CI [0.13, 0.40], *p* < .001). **Fig. 4A** illustrates the raw probability distributions for these trial-history effects. Given the unbalanced trial counts inherent to performance-dependent measures (e.g., fewer post-error trials, n = 9,874, compared to post-correct trials, n = 53,829), we based subsequent interaction analyses on estimated marginal means to ensure appropriate weighting.

The Task × Prev_Error × Stimulus_Switch interaction (OR = 4.14, 95%CI [1.42, 13.82], *p* = .013; **Fig. 4B**) revealed that in the pro-saccade task, switching produced lower error rates than repetition following both correct and error trials (*p*s < .001). In the anti-saccade task, switching reduced error rates following correct trials (*p* = .01) but increased them following errors (*p* < .001). Critically, stimulus repetition led to consistently lower error probabilities following error compared to correct trials across both tasks (*p*s < .001). In contrast, switching stimuli showed no significant difference between post-error and post-correct trials (*p*s > .34). These findings demonstrate that an overall switch advantage under simple conditions gives way to a repetition benefit following errors.

The Treatment × Task × Prev_Error interaction (OR = 1.519, 95%CI [1.007, 2.291], *p* = .046; **Fig 4C**) showed enhanced post-error adjustment under LT, with lower error probability in the LT group than the PLC group following error trials in both tasks (*p*s < .01), no group differences following correct trials (*p*s > .57).

The Treatment × Task × Stimulus Switch interaction (OR = 1.52, 95%CI [1.02, 2.30], *p* = .047; **Fig 4D**) revealed that in the anti-saccade task, the LT group demonstrated lower error rates than the PLC group during stimulus repetition trials (*p* = .02), while no treatment difference was found during switching trials (*p* = .54) or in the pro-saccade task (*p*s > .13).

Treatment effects also varied among sex, as revealed by the Treatment × Sex interaction (OR = 1.31, 95%CI [1.12, 1.54], *p* < .001) and Treatment × Sex × Prev_Error interaction (OR = 1.386, 95%CI [1.024, 1.875], *p* = .034; **Fig 4E**). LT females showed consistently lower error probability than PLC females (*p*s < .001). In contrast, LT males showed higher error probability than PLC males following correct trials (*p* = .001) but comparable performance following error trials (*p* = .88). While males showed lower error probability than females in the PLC group (*p* < .001), this sex difference was not present in the LT group (*p* = .79).

**Fig 4.**
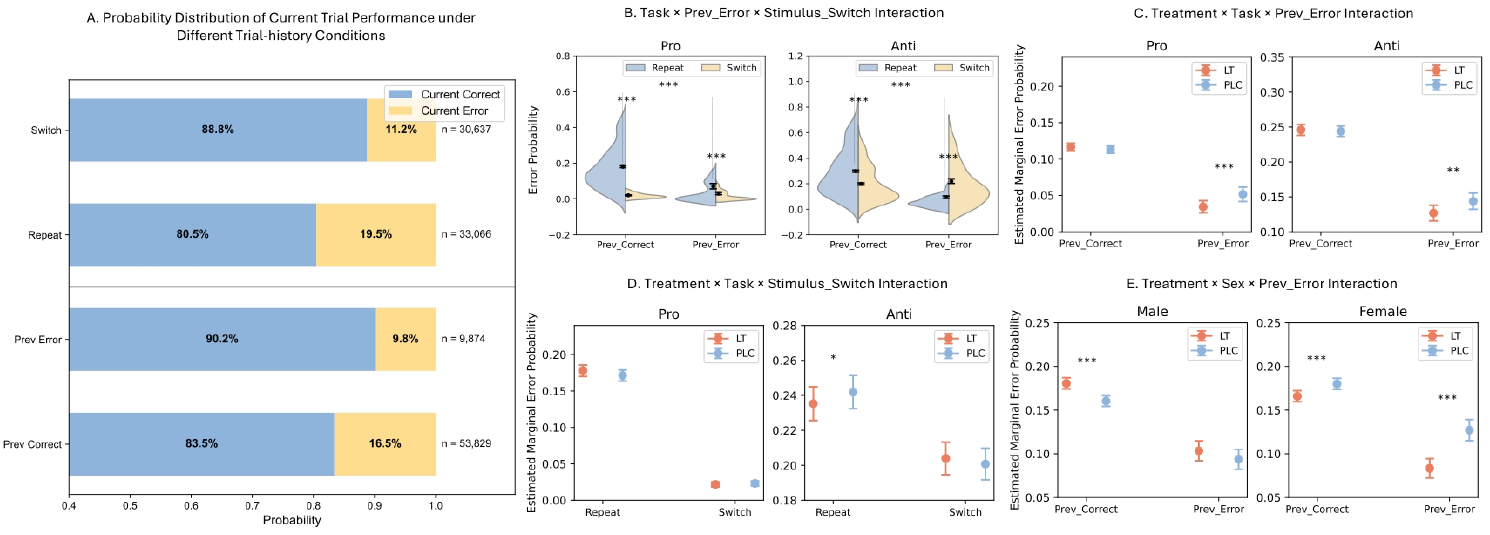
Dynamic modulation of current trial error probability by trial history. **(A)** Probability distributions of current trial performance categorized by switch versus repeat trials (upper panel) and post-error versus post- correct trials (lower panel). (B-E) Interaction effects on error probability for: **(B)** Task × Prev_Error × Stimulus Switch; **(C)** Treatment × Task × Prev_Error; **(D)** Treatment × Task × Stimulus Switch; and **(E)** Treatment × Sex × Prev_Error. **Note:** Violin plots depict the raw data distribution. Error bars represent 95% confidence intervals of the estimated marginal means (EMMeans) derived from generalized linear mixed models, accounting for unbalanced trial counts between conditions. ^*^*p*<.05, ^**^*p*<.01, ^***^*p*<.001.

#### 3.3.2 Current-trial Latency Adjustment

Significant main effects emerged for Prev_Error (*β* = -151.65, SE = 6.47, 95% CI [-164.33, - 138.97], *p* < .001) and Stimulus_Switch (*β* = -224.81, SE = 2.53, 95% CI [-229.77, -219.85], *p* < .001). Current trial latencies were shorter following errors compared to correct trials (291 ms vs. 353 ms) and following stimulus switches compared to repeats (228 ms vs. 416 ms; for distributions, see **Fig. 5A**).

The Task × Prev_Error × Stimulus_Switch interaction (*β* = -82.53, SE = 10.10, 95% CI [- 102.33, -62.73], *p* < .001; **Fig. 5B**) revealed that in pro-saccade task, responses following errors showed shorter latencies than those following correct trials during stimulus repetition (*p* < .001), but this difference was not observed during stimulus switching (*p* = .44). In anti-saccade task, responses following errors consistently showed shorter latencies than those following correct trials across both repeat and switch conditions (*p*s = .001).

The Treatment × Sex × Task interaction (*β* = -26.26, SE = 5.25, 95% CI [-36.54, -15.98], *p* < .001; **Fig. 5C**) showed that LT reduced latencies in females specifically during pro-saccade tasks (vs. PLC: *p* = .004), with no effects in anti-saccade tasks (*p* = .21) or in males (*p*s > .54). Sex comparisons showed that PLC males demonstrated consistently shorter latencies than PLC females across both tasks (*p*s < .001). Under LT, this male advantage persisted in anti-saccade tasks (*p* = .002) but disappeared in pro-saccade tasks (*p* = .33).

The Treatment × Sex × Stimulus_Switch interaction (*β* = -14.04, SE = 5.03, 95% CI [-23.89, - 4.19], *p* = .003; **Fig. 5D**) revealed that LT females exhibited shorter latencies than PLC females during stimulus repetitions (436ms vs. 440ms, *p* = .01) but longer latencies during switches (255ms vs. 250ms, *p* = .02). Males showed no treatment effects (*p*s > .75) and demonstrated consistently shorter latencies than females across all conditions (*p*s < .05).

**Fig 5.**
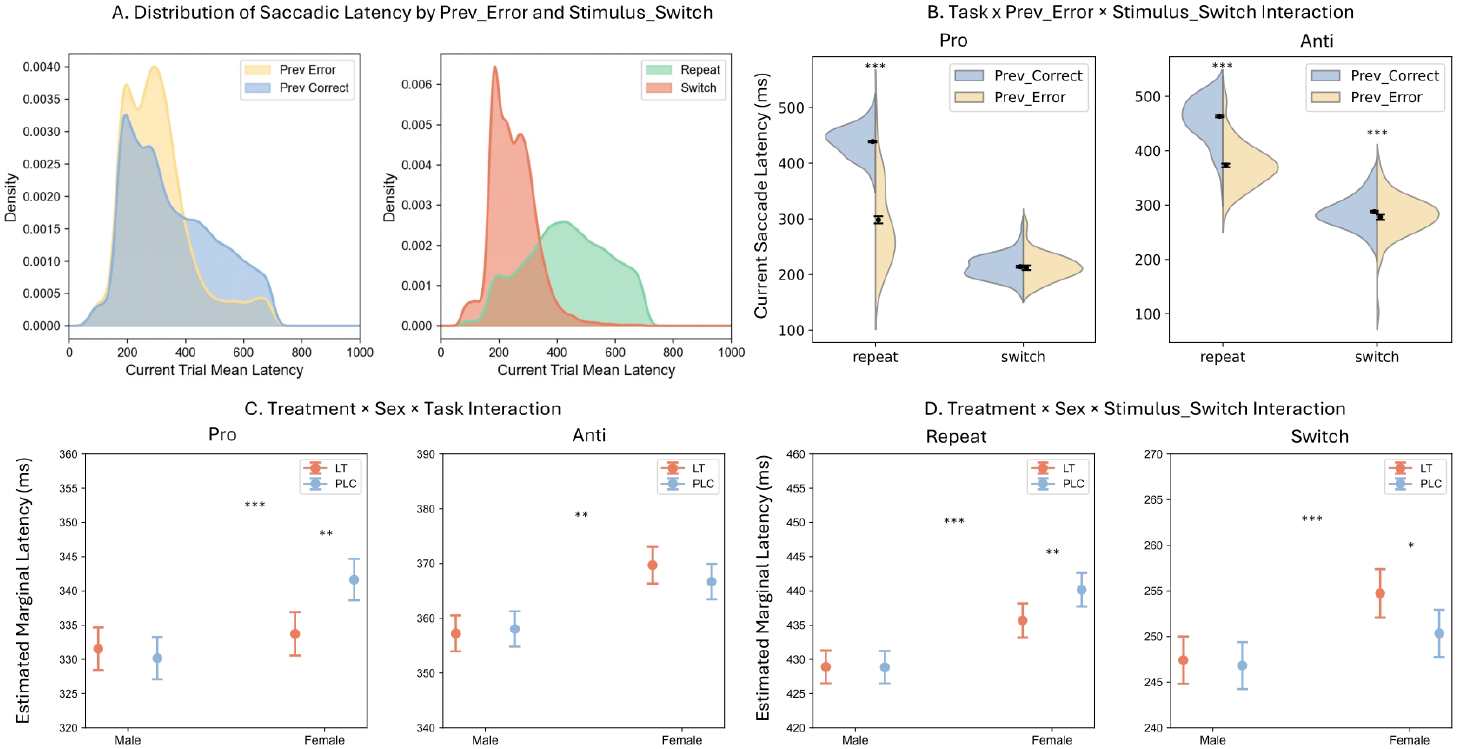
Dynamic modulation of saccadic latency by trial history. **(A)** Kernel density estimation (KDE) of raw saccadic latency distributions. Note the peaked distributions for post-error (left) and switch (right) trials compared to their counterparts. **(B–D)** Three-way interaction effects on saccadic latency for: **(B)** Task × Prev_Error × Stimulus Switch; **(C)** Treatment × Sex × Task; and **(D)** Treatment × Sex × Stimulus Switch. **Note:** Violin plots in (B) depict raw data distributions. Error bars represent 95% confidence intervals of the estimated marginal means (EMMs) derived from Linear Mixed Models (LMM), accounting for unbalanced trial counts. ^*^ *p* < .05, ^**^ *p* < .01, ^***^ *p* < .001.

## 4 Discussion

Our pharmacological eye-tracking study employed an emotional anti-saccade task to demonstrate an anxiolytic potential of transient angiotensin II receptor blockade via losartan and showed that it modulates both top-down goal-directed attention and bottom-up stimulus-driven processing with partially sex-dependent effects. By integrating analyses of initial oculomotor responses with trial-history informed dynamic control (TIDC), we demonstrate that losartan induces a sexually dimorphic reconfiguration of attentional processes, with findings highlighting that:

1. Independent of treatment, we observed a general “post-error repetition benefit” where performance improved on repetition trials following errors, and a “task-dependent switch effect”, whereas switching enhanced performance in the pro-saccade task regardless of prior outcome, which together led to improved performance in the anti-saccade specifically after correct trials yet impaired performance following errors.
2. Losartan enhanced the general utilization of contextual information by reducing error probabilities on both post-error and stimulus repetition trials.
3. Most critically, losartan reconfigured attentional processing in a sex-divergent manner. In females, it induced a shift that refined motor execution (reduced endpoint error) and enhanced flexibility (e.g., adaptive latency adjustment). In males, losartan induced a induced motor precision deficits (increased endpoint error) and reduced cognitive flexibility (e.g., elevated post-correct errors). This may reflect a failure to disengage from effortful control when it was not beneficial.

### 4.1 Context-Sensitive Adaptive Control

Our trial-history informed dynamic control (TIDC) analysis revealed a core principle of adaptive control: context-sensitive arbitration between cognitive stability and flexibility.

First, we observed a robust “post-error repetition benefit”—higher accuracy on repetition trials following errors. This reflects a strategic shift toward cognitive stability after failures, a process thought to rely on fronto-cingulate circuits. Specifically, the anterior cingulate cortex (ACC) detects performance errors and signals the need for increased top-down control to stabilize behavior and prevent consecutive errors (Botvinick et al., 2004; Danielmeier & Ullsperger, 2011).

Second, we found a “task-dependent switch effect”, demonstrating a nuanced gating of cognitive flexibility. In the low-demand pro-saccade task, switching conferred a general advantage. Conversely, in the cognitively demanding anti-saccade task which may require suppression of a prepotent response via dorsolateral prefrontal cortex (DLPFC) engagement (McDowell et al., 2008), the system strategically modulated flexibility: switching was beneficial after correct trials but detrimental after errors.

This pattern confirms that our paradigm effectively dissociated control sub-processes, showing that the system dynamically allocates resources by integrating current task demands with recent performance history (Musslick & Cohen, 2021; Shenhav et al., 2016) to optimize the stability-flexibility trade-off.

### 4.2 Baseline Performance in the PLC group

Analysis of baseline (PLC) performance revealed a sex difference in processing efficiency. Males generated significantly more precise initial saccades (i.e., smaller endpoint error) in the antisaccade task compared to females. As reduced endpoint error indicates that the first volitional eye movement landed closer to the intended target location, this suggests a male advantage in the planning and execution of the goal-directed motor command under baseline conditions.

This advantage extended to global performance metrics: within the TIDC framework, males in the PLC group also exhibited lower error probability and shorter latency than females. Crucially, this advantage in males remained stable and was not modulated by trial history, suggesting it it is based in differences in more automated execution efficiency rather than from context-sensitive, supervisory control (Hutton & Ettinger, 2006). Establishing these distinct baseline profiles is critical for interpreting the subsequent drug effects, as they indicate that males and females not only differed in performance level but also in their baseline modes of cognitive processing, representing divergent starting points for pharmacological intervention.

### 4.3 Effects of Losartan on Multidimensional Cognitive Control

Losartan reduced anxiety across sexes without concomitant changes in blood pressure. This dissociation between subjective state and peripheral cardiovascular activity likely reflects the delayed peak efficacy of losartan on hemodynamic function in normotensive subjects (Benicky et al., 2011; Xu et al., 2022), suggesting that the anxiolytic and subsequent cognitive effects are mediated centrally, independent of peripheral vascular changes (Salim & Jones, 2022).

Beyond subjective anxiety, losartan did not alter global inhibitory control, as indicated by comparable overall directional error rates between the LT and PLC groups. This suggests the drug does not non-specifically elevate basal inhibitory capacity. However, TIDC analysis revealed a significant enhancement in context-sensitive regulation: losartan reduced error probabilities conditional on trial history (e.g., post-error and repetition trials). This dissociation indicates that while losartan does not broadly suppress reflexive responses, it specifically improves the top-down monitoring system’s ability to utilize recent context to strategically prevent errors. This facilitation of context-dependent adaptive control implies an optimization of prefrontal networks and their interaction with subcortical affective and motor processing regions, including the anterior cingulate and lateral prefrontal cortex (Botvinick et al., 2004; Cools & D’Esposito, 2011; Zhuang et al., 2021). Importantly, this reconfiguration was sexually dimorphic, affecting both motor execution quality and adaptive control flexibility:

Regarding rapid sensorimotor processing, losartan exerted opposing sex-dependent effects. In females, the drug induced optimization, significantly decreasing endpoint error — suggesting a calibration of the initial motor command for enhanced precision. In males, it disrupted automatic processing, increasing endpoint errors and prolonging saccade latency, implying a cost of enhanced cognitive supervision on typically automatic subcortical-motor circuits.

Regarding dynamic adaptive control, these sex-divergent effects interacted with trial history. Following correct trials, when control demand is typically low, females on losartan maintained low error probabilities, effectively sustaining control. They also showed a strategic trade-off: faster responses on stimulus repeats and slower responses on switches, indicating adaptive resource allocation. In contrast, males on losartan showed elevated error probabilities after correct trials and lacked differential latency adjustments, reflecting a failure to disengage from the previous trial and more rigid behavioral adaptation.

The observed sex-divergent effects may be mechanistically grounded in the interaction between the RAS and dopaminergic neurotransmission. Angiotensin II via AT1 receptors is known to modulate dopamine release in the striatum and prefrontal cortex (Kobiec et al., 2021; Labandeira-Garcia et al., 2017). Given that cognitive control and flexibility rely on optimal dopamine levels following an ‘inverted-U’ shaped function (Cools & D’Esposito, 2011), the differential effects of losartan could reflect baseline differences in dopaminergic tone between sexes (Jacobs & D’Esposito, 2011). Specifically, if males operate near optimal dopaminergic levels at baseline, losartan-induced modulation might push them toward supra-optimal levels, resulting in the observed rigidity and motor deficits. Conversely, in females, who may exhibit different baseline dopaminergic dynamics possibly influenced by estrogen-RAS interactions (Leete et al., 2018; Nwia et al., 2023), AT1R blockade might optimize prefrontal-striatal signaling, thereby enhancing flexibility and precision.

In summary, our findings implicate the RAS in adaptive attention allocation alongside anxiolytic effects. Transient AT1R blockade induces a sex-specific reconfiguration of attentional processing: in females, it enhances both basic motor precision and higher-order adaptive control, while in males, it disrupts efficient automatic processing and reduces behavioral flexibility.

### 4.4 The Role of RAS in Attention and Clinical Implications

Our findings establish that AT1R blockade modulates core components of attentional control, providing direct evidence for a functional “brain-RAS” in cognition beyond its classical cardiovascular role. This underscores the RAS as a neuromodulatory system that shapes prefrontal-dependent processes such as contextual adaptation and error monitoring (Labandeira-García et al., 2014; Wright & Harding, 2010; Xu et al., 2025).

The robust sex-divergent effects indicate that the RAS influences cognition through interaction with sex-specific neuroendocrine milieus, likely involving estrogen-dopamine crosstalk. This necessitates the consideration of sex as a critical biological variable in both future research and clinical translation. Specifically, the cognitive optimization observed in females suggests AT1R antagonists may hold particular therapeutic promise for disorders where attentional control and affective regulation are impaired, such as anxiety disorders or ADHD, potentially offering a dual benefit for hypertensive patients with comorbid cognitive symptoms (Carnovale et al., 2023; Saavedra, 2012).

Methodologically, the study demonstrates that a trial-history informed approach is essential for detecting the nuanced, dynamic effects of neuromodulation on cognitive control, effects that are masked in conventional trial-averaged analyses. The use of emotional stimuli further bridges affective and cognitive domains, offering a mechanistic lens through which the anxiolytic properties of RAS blockers may be understood.

### 4.5 Limitations

The interpretation of our results is tempered by several limitations. First, the single-dose design precludes conclusions regarding the effects of chronic AT1R blockade, which may differ in both magnitude and mechanism. Second, while we report sex-dependent effects, the lack of hormonal profiling in female participants limits our ability to attribute these differences specifically to estrogenic modulation of the RAS. Third, while social (faces) and non-social (ovals) stimuli were matched for overall luminance and color, they inherently differed in low-level visual features (e.g., spatial frequency, contrast distribution, and shape complexity). These perceptual differences may contribute to the latency effects observed specifically in the non-social condition, and thus caution is warranted when interpreting the effects as purely reflective of social versus non-social cognitive processing. Fourth, our TIDC analysis was restricted to emotional face blocks due to the study design; thus, it remains unclear whether the enhanced context-sensitive regulation generalizes to non-social contexts.

Finally, while adequately powered for primary outcomes, the sample size may be insufficient to fully resolve complex higher-order interactions. Future studies should employ longer dosing regimens, control for female hormonal status, utilize perceptually matched stimuli, and extend TIDC modeling to non-affective tasks. Research in clinical populations will be crucial for establishing the translational validity of RAS modulation for cognitive enhancement.

In summary, this study positions the RAS as a key modulator of attentional control, with effects critically shaped by sex. The ability of acute AT1R blockade to enhance specific components of cognitive control, particularly in females, provides a foundation for developing targeted therapeutic strategies for neuropsychiatric disorders characterized by cognitive and affective dysregulation.

## Data availability

All relevant data will be made publicly available through the Open Science Framework (OSF) upon acceptance of the manuscript.

## Author Contributions

X.X. designed the experiment and provided the E-prime program. Q.Z. consulted on the experimental design. D.L. contributed to preliminary program debugging. J.W. managed drug administration and allocation. F.S. was responsible for experimental safety. M.H., K.F., and W.D. collected the data. M.H. performed the analysis and wrote the paper. T.X., W.C.C., and B.B. provided guidance on Losartan and the implementation of the double-blind procedure. B.B. supervised the entire project. All authors contributed to manuscript revision, approved the final version, and agree to be accountable for all aspects of the work.

## Funding

The project has been supported by the Health and Medical Research Fund Hong Kong (HMRF, 23243971), Ministry of Science and Technology of China (STI 2030 – Major Projects 2022ZD0208500), the University of Hong Kong seed funding and start-up schemes (2407102536; 2402101713).

## Competing Interests

The authors have nothing to disclose.

## Notes

### Competing Interest Statement

The authors have declared no competing interest.

### Clinical Trial

NCT06329050

### Author Declarations

Ethics committee of University of Electronic Science and Technology of China Ethics Committee gave ethical approval for this work (approval number: 106142024050929462)

### Summary of Updates

The author list has been updated to include Prof. Dezhong Yao. This addition correctly reflects his role as the principal investigator of the funding source (STI 2030 Major Projects 2022ZD0208500) already listed in the manuscript, as well as his contribution to study supervision.

## Reference

Adolphs, R. (2009). The social brain: Neural basis of social knowledge. Annual Review of Psychology, 60, 693–716. 10.1146/annurev.psych.60.110707.163514

Anderson, B. A. (2021). An adaptive view of attentional control. American Psychologist, 76(9), 1410–1422. 10.1037/amp0000917

Antoniades, C., Ettinger, U., Gaymard, B., Gilchrist, I., Kristjánsson, A., Kennard, C., John Leigh, R., Noorani, I., Pouget, P., Smyrnis, N., Tarnowski, A., Zee, D. S., & Carpenter, R. H. S. (2013). An internationally standardised antisaccade protocol. Vision Research, 84, 1–5. 10.1016/j.visres.2013.02.007

Bachmann, J., Feldmer, M., Ganten, U., Stock, G., & Ganten, D. (1991). Sexual dimorphism of blood pressure: Possible role of the renin-angiotensin system. The Journal of Steroid Biochemistry and Molecular Biology, 40(4), 511-IN2. 10.1016/0960-0760(91)90270-F

Bandelow, B., Michaelis, S., & Wedekind, D. (2017). Treatment of anxiety disorders. Dialogues in Clinical Neuroscience, 19(2), 93–107. 10.31887/DCNS.2017.19.2/bbandelow

Barr, D. J., Levy, R., Scheepers, C., & Tily, H. J. (2013). Random effects structure for confirmatory hypothesis testing: Keep it maximal. Journal of Memory and Language, 68(3). 10.1016/j.jml.2012.11.001

Benicky, J., Sánchez-Lemus, E., Honda, M., Pang, T., Orecna, M., Wang, J., Leng, Y., Chuang, D.-M., & Saavedra, J. M. (2011). Angiotensin II AT1 receptor blockade ameliorates brain inflammation. Neuropsychopharmacology: Official Publication of the American College of Neuropsychopharmacology, 36(4), 857–870. 10.1038/npp.2010.225

Boal, H. L., Christensen, B. K., & Goodhew, S. C. (2018). Social anxiety and attentional biases: A top-down contribution? Attention, Perception, & Psychophysics, 80(1), 42–53. 10.3758/s13414-017-1415-5

Bold K., & L M. (1999). Estrogen, natriuretic peptides and the renin–angiotensin system. Cardiovascular Research, 41(3), 524–531. 10.1016/S0008-6363(98)00324-1

Botvinick, M. M., Cohen, J. D., & Carter, C. S. (2004). Conflict monitoring and anterior cingulate cortex: An update. Trends in Cognitive Sciences, 8(12), 539–546. 10.1016/j.tics.2004.10.003

Brauer, M., & Curtin, J. J. (2018). Linear mixed-effects models and the analysis of nonindependent data: A unified framework to analyze categorical and continuous independent variables that vary within-subjects and/or within-items. Psychological Methods, 23(3), 389–411. 10.1037/met0000159

Capron, D. W., Norr, A. M., Allan, N. P., & Schmidt, N. B. (2017). Combined “top-down” and “bottom-up” intervention for anxiety sensitivity: Pilot randomized trial testing the additive effect of interpretation bias modification. Journal of Psychiatric Research, 85, 75–82. 10.1016/j.jpsychires.2016.11.003

Carnovale, C., Perrotta, C., Baldelli, S., Cattaneo, D., Montrasio, C., Barbieri, S. S., Pompilio, G., Vantaggiato, C., Clementi, E., & Pozzi, M. (2023). Antihypertensive drugs and brain function: Mechanisms underlying therapeutically beneficial and harmful neuropsychiatric effects. Cardiovascular Research, 119(3), 647–667. 10.1093/cvr/cvac110

Chen, N. T. M., Clarke, P. J. F., Watson, T. L., MacLeod, C., & Guastella, A. J. (2014). Biased saccadic responses to emotional stimuli in anxiety: An antisaccade study. PLOS ONE, 9(2), e86474. 10.1371/journal.pone.0086474

Chen, Y. F., Naftilan, A. J., & Oparil, S. (1992). Androgen-dependent angiotensinogen and renin messenger RNA expression in hypertensive rats. Hypertension (Dallas, Tex.: 1979), 19(5), 456–463. 10.1161/01.hyp.19.5.456

Chrissobolis, S., Luu, A. N., Waldschmidt, R. A., Yoakum, M. E., & D’Souza, M. S. (2020). Targeting the renin angiotensin system for the treatment of anxiety and depression. Pharmacology, Biochemistry, and Behavior, 199, 173063. 10.1016/j.pbb.2020.173063

Cieslik, E. C., Seidler, I., Laird, A. R., Fox, P. T., & Eickhoff, S. B. (2016). Different involvement of subregions within dorsal premotor and medial frontal cortex for pro- and antisaccades. Neuroscience & Biobehavioral Reviews, 68, 256–269. 10.1016/j.neubiorev.2016.05.012

Collister, J. P., Soucheray, S. L., & Osborn, J. W. (2002). Chronic hypotensive effects of losartan are not dependent on the actions of angiotensin II at AT 2 receptors. Journal of Cardiovascular Pharmacology, 39(1), 107–116. 10.1097/00005344-200201000-00012

Cools, R., & D’Esposito, M. (2011). Inverted-U-shaped dopamine actions on human working memory and cognitive control. Biological Psychiatry, 69(12), e113–125. 10.1016/j.biopsych.2011.03.028

Danielmeier, C., & Ullsperger, M. (2011). Post-error adjustments. Frontiers in Psychology, 2. 10.3389/fpsyg.2011.00233

Gao, N., Wang, H., Xu, X., Yang, Z., & Zhang, T. (2021). Angiotensin II induces cognitive decline and anxiety-like behavior via disturbing pattern of theta-gamma oscillations. Brain Research Bulletin, 174, 84–91. 10.1016/j.brainresbull.2021.06.002

Heimberg, R. G., Horner, K. J., Juster, H. R., Safren, S. A., Brown, E. J., Schneier, F. R., & Liebowitz, M. R. (1999). Psychometric properties of the liebowitz social anxiety scale. Psychological Medicine, 29(1), 199–212. 10.1017/S0033291798007879

Hutton, S. B., & Ettinger, U. (2006). The antisaccade task as a research tool in psychopathology: A critical review. Psychophysiology, 43(3), 302–313. 10.1111/j.1469-8986.2006.00403.x

Jacobs, E., & D’Esposito, M. (2011). Estrogen shapes dopamine-dependent cognitive processes: Implications for women’s health. The Journal of Neuroscience: The Official Journal of the Society for Neuroscience, 31(14), 5286–5293. 10.1523/JNEUROSCI.6394-10.2011

Kanaan, D., & Moacdieh, N. M. (2022). Eye tracking to evaluate the effects of interruptions and workload in a complex task. Human Factors, 64(7), 1168–1180. 10.1177/0018720821990487

Kanwisher, N., McDermott, J., & Chun, M. M. (1997). The fusiform face area: A module in human extrastriate cortex specialized for face perception. The Journal of Neuroscience: The Official Journal of the Society for Neuroscience, 17(11), 4302–4311. 10.1523/JNEUROSCI.17-11-04302.1997

Kobiec, T., Otero-Losada, M., Chevalier, G., Udovin, L., Bordet, S., Menéndez-Maissonave, C., Capani, F., & Pérez-Lloret, S. (2021). The renin–angiotensin system modulates dopaminergic neurotransmission: A new player on the scene. Frontiers in Synaptic Neuroscience, 13. 10.3389/fnsyn.2021.638519

Labandeira-García, J. L., Garrido-Gil, P., Rodriguez-Pallares, J., Valenzuela, R., Borrajo, A., & Rodríguez-Perez, A. I. (2014). Brain renin-angiotensin system and dopaminergic cell vulnerability. Frontiers in Neuroanatomy, 8. 10.3389/fnana.2014.00067

Labandeira-Garcia, J. L., Rodríguez-Perez, A. I., Garrido-Gil, P., Rodriguez-Pallares, J., Lanciego, J. L., & Guerra, M. J. (2017). Brain renin-angiotensin system and microglial polarization: Implications for aging and neurodegeneration. Frontiers in Aging Neuroscience, 9, 129. 10.3389/fnagi.2017.00129

Leete, J., Gurley, S., & Layton, A. (2018). Modeling sex differences in the renin angiotensin system and the efficacy of antihypertensive therapies. Computers & Chemical Engineering, 112, 253–264. 10.1016/j.compchemeng.2018.02.009

Li, J., Yang, X., Zhou, F., Liu, C., Wei, Z., Xin, F., Daumann, B., Daumann, J., Kendrick, K. M., & Becker, B. (2020). Modafinil enhances cognitive, but not emotional conflict processing via enhanced inferior frontal gyrus activation and its communication with the dorsomedial prefrontal cortex. Neuropsychopharmacology: Official Publication of the American College of Neuropsychopharmacology, 45(6), 1026–1033. 10.1038/s41386-020-0625-z

Luna, B., Velanova, K., & Geier, C. F. (2008). Development of eye-movement control. Brain and Cognition, A Hundred Years of Eye Movement Research in Psychiatry, 68(3), 293–308. 10.1016/j.bandc.2008.08.019

McDowell, J. E., Dyckman, K. A., Austin, B. P., & Clementz, B. A. (2008). Neurophysiology and neuroanatomy of reflexive and volitional saccades: Evidence from studies of humans. Brain and Cognition, A Hundred Years of Eye Movement Research in Psychiatry, 68(3), 255–270. 10.1016/j.bandc.2008.08.016

Meteyard, L., & Davies, R. A. I. (2020). Best practice guidance for linear mixed-effects models in psychological science. Journal of Memory and Language, 112, 104092. 10.1016/j.jml.2020.104092

Monsell, S. (2003). Task switching. Trends in Cognitive Sciences, 7(3), 134–140. 10.1016/s1364-6613(03)00028-7

Munoz, D. P., & Everling, S. (2004). Look away: The anti-saccade task and the voluntary control of eye movement. Nature Reviews Neuroscience, 5(3), 218–228. 10.1038/nrn1345

Musslick, S., & Cohen, J. D. (2021). Rationalizing constraints on the capacity for cognitive control. Trends in Cognitive Sciences, 25(9), 757–775. 10.1016/j.tics.2021.06.001

Noorani, I., & Carpenter, R. H. S. (2016). The LATER model of reaction time and decision. Neuroscience & Biobehavioral Reviews, 64, 229–251. 10.1016/j.neubiorev.2016.02.018

Notebaert, W., Houtman, F., Opstal, F. V., Gevers, W., Fias, W., & Verguts, T. (2009). Post-error slowing: An orienting account. Cognition, 111(2), 275–279. 10.1016/j.cognition.2009.02.002

Nwia, S. M., Leite, A. P. O., Li, X. C., & Zhuo, J. L. (2023). Sex differences in the renin-angiotensin-aldosterone system and its roles in hypertension, cardiovascular, and kidney diseases. Frontiers in Cardiovascular Medicine, 10. 10.3389/fcvm.2023.1198090

Oberauer, K. (2024). The meaning of attention control. Psychological Review, 131(6), 1509–1526. 10.1037/rev0000514

Ott, T., & Nieder, A. (2019). Dopamine and cognitive control in prefrontal cortex. Trends in Cognitive Sciences, 23(3), 213–234. 10.1016/j.tics.2018.12.006

Pinto, Y., van der Leij, A. R., Sligte, I. G., Lamme, V. A. F., & Scholte, H. S. (2013). Bottom-up and top-down attention are independent. Journal of Vision, 13(3), 16. 10.1167/13.3.16

Prasad, D., Shkreli, L., De Giorgi, R., Costi, S., & Reinecke, A. (2025). Acute angiotensin receptor blockade and mnemonic discrimination in healthy participants. Journal of Affective Disorders, 375, 293–296. 10.1016/j.jad.2025.01.119

Reinecke, A., Browning, M., Klein Breteler, J., Kappelmann, N., Ressler, K. J., Harmer, C. J., & Craske, M. G. (2018). Angiotensin regulation of amygdala response to threat in high-trait-anxiety individuals. Biological Psychiatry. Cognitive Neuroscience and Neuroimaging, 3(10), 826–835. 10.1016/j.bpsc.2018.05.007

Saavedra J. M. (2012). Angiotensin II AT1 receptor blockers as treatments for inflammatory brain disorders. Clinical Science, 123(10), 567–590. 10.1042/CS20120078

Sacher, J., Neumann, J., Okon-Singer, H., Gotowiec, S., & Villringer, A. (2013). Sexual dimorphism in the human brain: Evidence from neuroimaging. Magnetic Resonance Imaging, 31(3), 366–375. 10.1016/j.mri.2012.06.007

Salim, H., & Jones, A. M. (2022). Angiotensin II receptor blockers (ARBs) and manufacturing contamination: A retrospective national register study into suspected associated adverse drug reactions. British Journal of Clinical Pharmacology, 88(11), 4812–4827. 10.1111/bcp.15411

Shenhav, A., Cohen, J. D., & Botvinick, M. M. (2016). Dorsal anterior cingulate cortex and the value of control. Nature Neuroscience, 19(10), 1286–1291. 10.1038/nn.4384

Si, Y., Wang, L., & Zhao, M. (2022). Anti-saccade as a tool to evaluate neurocognitive impairment in alcohol use disorder. Frontiers in Psychiatry, 13, 823848. 10.3389/fpsyt.2022.823848

Sica, D. A., Gehr, T. W. B., & Ghosh, S. (2005). Clinical pharmacokinetics of losartan. Clinical Pharmacokinetics, 44(8), 797–814. 10.2165/00003088-200544080-00003

Spielberger, C. D. (2012). State-trait anxiety inventory for adults [Dataset]. 10.1037/t06496-000

Sussman, T. J., Jin, J., & Mohanty, A. (2016). Top-down and bottom-up factors in threat-related perception and attention in anxiety. Biological Psychology, Determinants and Associations of Threat-Related Cognitive Biases: Cognitive and Neurophysiological Perspectives, 121, 160–172. 10.1016/j.biopsycho.2016.08.006

Swiercz, A. P., Iyer, L., Yu, Z., Edwards, A., Prashant, N. M., Nguyen, B. N., Horvath, A., & Marvar, P. J. (2020). Evaluation of an angiotensin type 1 receptor blocker on the reconsolidation of fear memory. Translational Psychiatry, 10(1), 363. 10.1038/s41398-020-01043-6

Watson, D., Clark, L. A., & Tellegen, A. (1988). Development and validation of brief measures of positive and negative affect: The PANAS scales. Journal of Personality and Social Psychology, 54(6), 1063–1070. 10.1037/0022-3514.54.6.1063

Wright, J. W., & Harding, J. W. (2010). The brain RAS and alzheimer’s disease. Experimental Neurology, 223(2), 326–333. 10.1016/j.expneurol.2009.09.012

Xu, T., Chen, Z., Zhou, X., Wang, L., Zhou, F., Yao, D., Zhou, B., & Becker, B. (2024). The central renin–angiotensin system: A genetic pathway, functional decoding, and selective target engagement characterization in humans. Proceedings of the National Academy of Sciences, 121(8), e2306936121. (world). 10.1073/pnas.2306936121

Xu, T., Fu, K., Yao, D., Ma, C., & Becker, B. (2025). The brain renin angiotensin system: A novel precision target for neurofunctional symptom regulation. OSF. 10.31234/osf.io/fbcv4_v1

Xu, T., Zhou, X., Jiao, G., Zeng, Y., Zhao, W., Li, J., Yu, F., Zhou, F., Yao, S., & Becker, B. (2022). Angiotensin antagonist inhibits preferential negative memory encoding via decreasing hippocampus activation and its coupling with the amygdala. Biological Psychiatry: Cognitive Neuroscience and Neuroimaging, 7(10), 970–978. 10.1016/j.bpsc.2022.05.007

Xu, T., Zhou, X., Kanen, J. W., Wang, L., Li, J., Chen, Z., Zhang, R., Jiao, G., Zhou, F., Zhao, W., Yao, S., & Becker, B. (2023). Angiotensin blockade enhances motivational reward learning via enhancing striatal prediction error signaling and frontostriatal communication. Molecular Psychiatry, 28(4), 1692–1702. 10.1038/s41380-023-02001-6

Xu, X., Li, J., Chen, Z., Kendrick, K. M., & Becker, B. (2019). Oxytocin reduces top-down control of attention by increasing bottom-up attention allocation to social but not non-social stimuli—A randomized controlled trial. Psychoneuroendocrinology, 108, 62–69. 10.1016/j.psyneuen.2019.06.004

Yang, C., He, L., Liu, Y., Lin, Z., Luo, L., & Gao, S. (2023). Anti-saccades reveal impaired attention control over negative social evaluation in individuals with depressive symptoms. Journal of Psychiatric Research, 165, 64–69. 10.1016/j.jpsychires.2023.07.016

Zhou, F., Geng, Y., Xin, F., Li, J., Feng, P., Liu, C., Zhao, W., Feng, T., Guastella, A. J., Ebstein, R. P., Kendrick, K. M., & Becker, B. (2019). Human extinction learning is accelerated by an angiotensin antagonist via ventromedial prefrontal cortex and its connections with basolateral amygdala. Biological Psychiatry, 86(12), 910–920. 10.1016/j.biopsych.2019.07.007

Zhou, X., Xu, T., Zeng, Y., Zhang, R., Qi, Z., Zhao, W., Kendrick, K. M., & Becker, B. (2023). The angiotensin antagonist losartan modulates social reward motivation and punishment sensitivity via modulating midbrain-striato-frontal circuits. The Journal of Neuroscience, 43(3), 472–483. 10.1523/JNEUROSCI.1114-22.2022

Zhuang, Q., Zheng, X., Becker, B., Lei, W., Xu, X., & Kendrick, K. M. (2021). Intranasal vasopressin like oxytocin increases social attention by influencing top-down control, but additionally enhances bottom-up control. Psychoneuroendocrinology, 133, 105412. 10.1016/j.psyneuen.2021.105412

Zhuang, Q., Zheng, X., Yao, S., Zhao, W., Becker, B., Xu, X., & Kendrick, K. M. (2022). Oral administration of oxytocin, like intranasal administration, decreases top-down social attention. International Journal of Neuropsychopharmacology, 25(11), 912–923. 10.1093/ijnp/pyac059

Zika, O., Appel, J., Klinge, C., Shkreli, L., Browning, M., Wiech, K., & Reinecke, A. (2024). Reduction of aversive learning rates in pavlovian conditioning by angiotensin II antagonist losartan: A randomized controlled trial. Biological Psychiatry, 96(4), 247–255. 10.1016/j.biopsych.2024.01.020

