## Supplementary material for "Angiotensin II Regulates Anxiety and Social-Affective Top-Down and Bottom-Up Attention Control in a Sex-dependent Manner": Complete Supplementary Materials (from another version of this preprint: version 1, 2025-10-28)

### Exclusion Criteria

- History of hepatic, renal, gastrointestinal, respiratory, endocrine, neuropsychiatric, cardiovascular diseases.
- A history of cardiac disease including arrhythmias; history of syncope or unexplained loss of consciousness; history of renal stones or renal failure; history of diabetes mellitus or diabetes insipidus.
- History of or present renal artery stenosis (reduced blood flow to the kidneys).
- History of hypersensitivity or allergic reaction to any medication or hormone, strong allergic reaction to food, or general tendency for allergies.
- Known hypersensitivity to losartan or related agents (e.g. telmisartan), antidiuretic hormone or any other component of the formulation; known hypersensitivity to any other angiotensin II receptor antagonist.
- Subjects with hypertension (BP > 140/90mmHg) or hypothension (BP < 90/60mmHg).
- Current hyponatremia (low levels of sodium in the blood) (symptoms include nausea, fatigue and weakness or swelling of hands, feet or legs)
- History of alcohol or drug abuse； smoker (≥ 10 cigarettes or ≥ 3 cigars or ≥ 3 pipes/day)
- Blood donation (≤ 1 month prior to administration)
- Take oral contraceptives or receive hormonal medications in the three months prior to the experiment.
- Pregnant or breastfeeding.
- Current disorder, including infections such as COVID, flu or cold.

### Pre-Screening Procedures

Prior to participation, all volunteers underwent an electrocardiogram (ECG) examination to rule out potential cardiovascular contraindications. To minimize potential confounds from psychoactive substances, participants were required to abstain from caffeine and alcohol for 24 hours prior to the testing session.

For female participants, a urine-based pregnancy test was administered on the day of the experiment to exclude pregnancy. Additionally, the phase of their menstrual cycle was documented based on self-report following established procedures (Gao et al., 2022; Garver-Apgar et al., 2008) to account for potential hormonal influences.

Based on these pre-screening procedures, which applied stricter, study-specific health criteria, 55 volunteers were excluded from participation. Among them, 42 were excluded due to abnormal or potentially abnormal ECG findings, 11 due to blood pressure outside the target range (systolic >130 or diastolic <80 mmHg), and 2 due to abnormal resting heart rate (<60 or >90 bpm).

### Randomization and Blinding

The allocation sequence was generated by a computer-based random number generator, using block randomization with randomly varying block sizes of 3 and 6 to ensure balanced group sizes.

To conceal the allocation sequence and ensure double-blinding, an independent researcher prepared all medications. All capsules were identical in appearance and packaged in containers labeled with a unique code according to the randomization sequence. The research staff involved in participant recruitment and treatment administration received only these pre-coded containers.

They had no access to the allocation list and remained fully blinded to the treatment group assignments. This system successfully ensured that group allocation was concealed until all data collection was complete and the database was locked.

All interventions were administered by trained staff following a standardized protocol in a laboratory at the University of Electronic Science and Technology of China (UESTC), Chengdu, China. This was a single-session laboratory study, and all primary and secondary outcomes were collected during the session. To monitor potential adverse effects, a follow-up text message was sent to participants 24 – 48 hours after the experiment. No delayed discomfort or adverse events were reported.

**Table S1. Demographics and questionnaire scores between treatment groups**

| Variables | LT | PLC | *p*-value |
|---|---|---|---|
| Age, Years | 21.92±2.28 | 22.26±2.52 | 0.51 |
| PANAS-Positive | 29.50±4.70 | 27.47±5.44 | 0.09 |
| PANAS-Negative | 15.39±5.14 | 16.39±5.14 | 0.46 |
| LSAS-Anxiety | 47.29±12.17 | 45.47±8.62 | 0.45 |
| LSAS-Avoidance | 43.84±11.93 | 43.87±10.21 | 0.96 |
| TAI | 37.66±7.63 | 39.69±6.29 | 0.15 |
| SAI-Baseline | 35.29±8.19 | 37.58±6.67 | 0.68 |
| SAI-Post | 34.21±8.29 | 37.81±8.16 | 0.02* |

Values are presented as mean ± SD.

PANAS, Positive and Negative Affect Schedule; LSAS, Liebowitz Social Anxiety Scale; TAI and SAI, Trait and State subscales of the State-Trait Anxiety Inventory; LT, losartan; PLC, placebo.

SAI-Post: This between-group difference is consistent with the significant Treatment × Time interaction in the mixed ANOVA (see main text).

**Table S2. Cardiovascular activity measures**

| Physiological Measures | Baseline | | Peak plasma (90min) | | Post experiment | | *p* value |
|---|---|---|---|---|---|---|---|
| | LT | PLC | LT | PLC | LT | PLC | |
| Systolic blood pressure | 114.79±0.92 | 112.21±0.89 | 112.21±1.16 | 113.35±1.13 | 114.04±1.78 | 115.01±1.15 | 0.61 |
| Diastolic blood pressure | 72.00±0.98 | 72.39±0.96 | 71.42±1.38 | 72.42±1.35 | 74.06±1.30 | 74.47±1.27 | 0.77 |
| Heart rate | 73.78±1.48 | 74.22±1.45 | 69.86±1.56 | 70.38±1.52 | 69.68±1.57 | 69.66±1.53 | 0.89 |

Values are presented as mean ± SD.

*F* tests for the interaction of Treatment (Placebo, Losartan) × Sex (Male, Female) × Time (Baseline, Peak plasma (90min), Post experiment).

### Direction Error Rate

A 2 (Treatment: LT, PLC) × 2 (Sex: male, female) × 2 (Task: pro-saccade, anti-saccade) × 2 (Condition: social, non-social) mixed ANOVA revealed a significant main effect of Task ($F$ = 130.92, $p < .001$, $\eta^{2p}$ = 0.636), with higher error rates in the anti-saccade task (17.5%) compared to the pro-saccade task (1.87%). A significant main effect of Condition was also observed ($F$ = 9.48, $p$ = .003, $\eta^{2p}$ = 0.112), with higher error rates in the social condition (11.22%) compared to the non-social condition (8.18%).

The analysis also revealed a significant Task × Condition interaction ($F$ = 6.29, $p$ = .014, $\eta^{2p}$ = 0.077). Simple effects analysis showed significantly higher error rates in the social condition compared to the non-social condition during the anti-saccade task (p = .006), while during the pro-saccade task, the difference between social and non-social conditions showed a marginal trend (p = .071).

To further examine the effects of different stimulus types, we conducted a 2 (Treatment: LT, PLC) × 2 (Sex: male, female) × 2 (Task: pro-saccade, anti-saccade) × 6 (Stimulus: shape, neutral, happy, fear, sad, angry) mixed ANOVA. Results showed a significant main effect of Stimulus ($F$ = 4.04, $p$ = .003, $\eta^{2p}$ = 0.222). Post-hoc comparisons revealed that shapes elicited significantly lower error rates (8.18%) compared to all emotional stimulus ($p$s < .05). Fear induced the highest error rates (21.92%), which were significantly higher than shapes, neutral, happy, and angry ($p$s < .05), but not significantly different from sad ($p$ = .25).

A significant Task × Stimulus interaction was also found ($F$ = 5.08, $p < .001$, $\eta^{2p}$ = 0.264). Simple effects analysis revealed that in the pro-saccade task, shapes only showed significantly lower error rates compared to neutral stimuli ($p$ = .036), with no significant differences from other stimulus ($p$s > .13). In the anti-saccade task, shapes showed significantly lower error rates compared to all emotional faces (ps < .05); neutral showed significantly lower error rates compared to fear and sad ($p$s < .05); happy showed significantly lower error rates compared to fear($p$ = .05) and fear exhibited significantly higher error rates compared to all other conditions except sad faces ($p$s < .05), demonstrating the strongest interference with attention control. Taken together, these findings indicate that the interference effect of fear was particularly pronounced in the anti- rather than in the pro-saccade task.

### Endpoint error of first correct saccade

For successfully executed first saccades, a 2 (Treatment: LT, PLC) × 2 (Sex: male, female) × 2 (Task: pro-saccade, anti-saccade) × 2 (Condition: social, non-social) mixed ANOVA revealed a significant main effect of Task ($F$ = 85.67, $p < .001$, $\eta^{2p}$ = 0.537), with larger endpoint error in the anti-saccade task compared to the pro-saccade task (123.86px vs 65.04px). A significant main effect of Condition was also observed ($F$ = 8.44, $p$ = .005, $\eta^{2p}$ = 0.102), with larger endpoint error in the non-social condition compared to the social condition (99.16px vs 89.74px).

To further examine the effects of different stimulus types, we conducted a 2 (Treatment: LT, PLC) × 2 (Sex: male, female) × 2 (Task: pro-saccade, anti-saccade) × 6 (Stimulus: shape, neutral, happy, fear, sad, angry) mixed ANOVA. Results showed a significant main effect of Stimulus ($F$ = 2.83, $p$ = .022, $\eta^{2p}$ = 0.168). Post-hoc comparisons revealed that shapes elicited significantly larger endpoint error (99.16px) compared to all emotional stimulus ($p$s < .05), while no significant differences were found among other emotional stimulus ($p$s > .10). No significant Task × Stimulus interaction ($p$ = .39) or other meaningful effects were observed.

### Latency of first correct saccade

A 2 (Treatment: LT, PLC) × 2 (Sex: male, female) × 2 (Task: pro-saccade, anti-saccade) × 2 (Condition: social, non-social) mixed ANOVA revealed a significant main effect of Task ($F$ = 719.81, $p < .001$, $\eta^{2p}$ = 0.907), with higher latency in the anti-saccade task compared to the pro-saccade task (302.80ms vs 225.56ms). A significant main effect of Condition was also observed ($F$ = 108.02, $p < .001$, $\eta^{2p}$ = 0.593), with higher latency in the non-social condition compared to the social condition (275.05ms vs 253.31ms).

To further examine the effects of different stimulus types, we conducted a 2 (Treatment: LT, PLC) × 2 (Sex: male, female) × 2 (Task: pro-saccade, anti-saccade) × 6 (Stimulus: shape, neutral, happy, fear, sad, angry) mixed ANOVA. Results showed a significant main effect of Stimulus ($F$ = 28.91, $p < .001$, $\eta^{2p}$ = 0.674). Post-hoc comparisons revealed that shapes elicited significantly higher latency compared to all emotional stimulus ($p$s < .001), while no significant differences were found among emotional stimulus ($p$s > .11).
